# Advanced imaging to improve remission rates in patients undergoing transsphenoidal surgery for Cushing’s disease

**DOI:** 10.1101/2020.07.04.20146498

**Authors:** Chan Hee Koh, Danyal Z. Khan, Hugo Layard Horsfall, Ahmad Ali, Ronneil Digpal, Jane Evanson, Hani J. Marcus, Márta Korbonits

## Abstract

**INTRODUCTION:** Cushing’s disease is a condition of hypercortisolaemia caused by a functional adrenocorti-cotropic hormone secreting pituitary adenoma. Surgical management is the first line of treatment for Cushing’s disease, but remains challenging in some cases. Various imaging modalities have been tried in an attempt to improve surgical remission rates. However, the sizes and certainties of any effect, in comparison to standard preoperative MRI imaging, remain unclear.

**METHODS:** This protocol has been developed according to the PRISMA-P 2015 standards. Searches of PubMed and Embase were conducted on 21 May 2020, and abstract screening commenced. The eligibility criteria, items for data extraction, and the analysis plan are laid out in this protocol, and uploaded prior to commencement of formal data extraction.

**DISCUSSION:** The results of this review will inform decision-making and priority-setting at the clinical, organisational and research levels. The results will be disseminated throughout the international neurosurgical community through presentations and publications.

## INTRODUCTION

Cushing’s disease is a condition where a functional adrenocorticotropic hormone (ACTH) releasing pituitary adenoma leads to secondary hypercortisolaemia. Untreated hypercortisolaemia produces a spectrum of symptomatology and signs which can lead to major morbidity (cardio- and cerebro-vascular disease, immunosuppression, osteoporosis amongst the many) with a consequent impact on lifespan ^1^.

Diagnosis of Cushing’s disease requires a carefully considered multidisciplinary approach with the involvement of endocrinology, radiology, interventional radiology and neurosurgery. The diagnosis of Cushing’s disease can be one of the most difficult challenges in endocrinology, as there are several complicating factors during the diagnostic process. Although Cushing’s disease is the most likely cause of increased ACTH secretion (and of endogenous hypercortisolaemia in general), ectopic sources of ACTH are possible ^2^.

The first line treatment for Cushing’s disease is surgical resection of the adenoma. A well targeted removal of the lesion preserves normal pituitary function whilst removing the source of ACTH ^1^. However, challenges remain in localisation of microadenomas and maximal safe resection.

Pre-operative MRIs are a well established feature in the diagnostic pipeline for Cushing’s disease. However, ACTH adenomas are often small at the time of presentation, and an estimated 40-60% are undetectable on preoperative MRI ^1;2;3^. Inferior petrosal sinus sampling (IPSS) can be used to confirm a pituitary source of ACTH secretion. Whether it helps to lateralise an otherwise undetectable adenoma is questionable, due to the often central location of the lesion and venous asymmetry ^1^. Several other imaging modalities have been tried in an attempt to improve remission rates ^3;4;5^. However, the sizes and the certainties of their effects on surgical remission rates for Cushing’s disease remains unclear.

We have set out to conduct a systematic review on the topic of imaging in Cushing’s disease to investigate this problem.

### OBJECTIVES

1. Estimate remission rates for Cushing’s disease using standard pre-operative MRI.
2. To identify other imaging modalities (e.g. intraoperative MRI, intraoperative ultrasound, PET) that have been used and their impact, if any, on remission rates.

### PROTOCOL

#### DEVELOPMENT

The development of this protocol adheres to the standards set out in the PRISMA-P 2015 statement ^6^.

#### REVIEW QUESTION

What is the effect of advanced imaging modalities (***intervention***), in comparison to standard preoperative MRI (***control***), on remission rates (***outcome***) in patients undergoing transsphenoidal surgery for Cushing’s disease (***population***)?

For the purposes of this review, standard preoperative MRI is defined as T1 and T2 sequences with and without contrast, with strengths up to 3-Teslas. Everything else (e.g. intraoperative MRI, dynamic contrast MRI, intraoperative ultrasound) is defined as advanced imaging ^7;8;9^.

A search strategy was developed systematically using the PICO approach based on the review question as outlined above (Table 1).

**Table 1.** Review question and search strategy using the PICO approach.

| PICO | Description | Search Terms |
| --- | --- | --- |
| **Population** | Cushing's Disease | (Cushing OR Cushing* OR ACTH OR ACTHoma OR ACTH* OR hypercortisolism OR hypercortisol*) |
|  | Transsphenoidal Surgery | (surgery OR surgical OR surg* OR operation OR operative OR transsphenoidal OR transsphenoid* OR endonasal OR endoscopic OR endoscop*) |
| **Intervention** | Advanced Imaging | (IPSS OR "inferior petrosal sinus sampling" OR imaging OR MRI OR ultrasound OR CT OR PET OR "positron emission tomography" OR hyperspectral OR HSI OR fluorescence OR spectrometry OR spectroscopy) |
| **Control** | Standard preoperative MRI | *No search* |
| **Outcome** | Remission | (localise OR localize OR lateralise OR lateralize OR outcome OR remission OR cure OR curative) |

### ELIGIBILITY CRITERIA

#### Exclusion Criteria

1. Non-primary study
2. Non-clinical study
3. Not English
4. No full-text
5. Single case-reports
6. No (active) Cushing’s disease
7. No surgical management
8. Case series of <10 patients
  a. Any papers excluded based on this criterion will be rescreened during analysis to pick up papers using novel/non-standard imaging techniques

9 No pre-operative or intraoperative imaging
10 No remission, reoperation or recurrence outcomes
11 Duplicate data
12 Other, please comment

#### Inclusion Criteria

1. Patients with Cushing’s disease undergoing transsphenoidal surgery
2. Pre-operative or intraoperative imaging
3. Reports surgical remission rates
4. Case series of >=10 patients that underwent similar treatment

#### SCREENING

##### Abstract Screening

Duplicates were removed by the first author (CHK). All identified abstracts are then to be screened by at least two independent reviewers. Conflicts will be settled through discussion.

Rayyan (www.rayyan.qcri.org; Qatar Computing Research Institute, Qatar), a free web application facilitating screening of abstracts, will be used during this stage ^10^.

##### Full Text Screening

Full-texts will be screened for eligibility concurrently to data collection. Both steps will utilise an in-house tool developed on the Google scripts platform (Google, California, USA) ^11^.

### DATA EXTRACTION

#### Outcomes

##### Primary Outcomes

- Remission after first surgery

##### Secondary Outcomes

- Overall remission
- Re-operation rate
- Recurrence rate

##### Comparative Outcomes

- e.g. relative risk, odds ratios etc. where available

##### Study Characteristics

###### Geography

- Region
- Country
- Center

###### Time

- Overall remission
- Re-operation rate
- Recurrence rate

###### Others

- Previous or concurrent treatments
- Definition of remission
- Surgical approach

##### Group Characteristics

###### Demographics

- Age
- Proportion of Males

###### Disease

- Size of adenoma
- Biochemical characteristics

###### Co-treatments

- Previous surgery
- Previous radiosurgery
- Previous medical therapy
- Adjuvant radiotherapy

###### Others

- Average follow up time
- Number lost to follow up

##### Imaging Characteristics

###### Preoperative MRI

- Strength of magnetic field (Tesla)
- Sequences

###### Other imaging modalities

- e.g. intraoperative MRI, intraoperative ultrasound, PET, SPECT

### STUDY QUALITY

Study quality and the risk of bias to be assessed using Cochrane’s RoB-2^12^, or ROBINS-1^13^ for randomised controlled trials or observational studies, respectively. These will be assessed at the study level.

To be presented as study quality dot plots, and robustness of analysis to quality will be tested with sensitivity analysis.

### ANALYSIS

Data synthesis will be in three parts:

1. Descriptive summary of study characteristics
2. Narrative synthesis of primary and secondary outcomes
3. Numerical synthesis of outcomes, with the imaging modality as dependent variable, and making adjustments for study-level confounders

We expect a very small number, if any, of controlled trials making direct comparisons between standard preoperative MRI and other imaging modalities. Therefore, meta-analysis of comparative outcomes (RR, OR, HR etc.) is not likely to be feasible.

On the other hand, we expect a far larger number of case series. We expect that calculating a global adjusted average to be feasible for surgical remission rates with standard preoperative MRI only. With this, it may be possible to compare results (either from single studies or a pool of studies) from a non-standard imaging modality to this global average.

Any papers with less than 10 patients, but reporting on a non-standard imaging technique, will be included during sensitivity analysis, and will be reported for exploratory purposes rather than for definitive conclusions.

The factors that will be investigated for confounding are outlined in the data extraction section above.

### Detailed meta-analysis plan

The appropriateness of numerical analysis to be judged during data extraction, considering the following factors:

- Numerical results and summary statistics reported
- Studies sufficiently similar to be pooled, or reasonable statistical adjustments can be made
- Sufficient number of studies to give a meaningful result (>5)
- No strong evidence of publication bias

If numerical analysis to be performed, standard analysis and statistics to be calculated and reported:

- Pooled ratios
- Estimate, 95% CI, 95% PI, p-value
- I^2^ measure of heterogeneity, with p-values

Publication bias to be assessed by Egger’s regression, visual inspection of funnel plot and Duval’s trim-and-fill.

Subgroup/meta-regression/multilevel analyses for investigation of heterogeneity

- MRI subtypes - e.g. Tesla, MTI sequences
- Tumour size - e.g. macroadenoma, microadenoma, radiologically undetectable adenoma

Meta-regression analysis if input variables are continuous

Sensitivity analysis - study design (RCT, cohort, case series), study quality and risk of bias

### STRENGTH OF EVIDENCE

To be summarised using the GRADE approach ^14^.

## DISCUSSION

At the time of writing, piloting of the study process had been completed, and full searches conducted on 21 May 2020. Single screen of abstract has been completed and awaiting completion of double screen. Formal data extraction not yet commenced.

The main objective of this review is to investigate the extent that the modality of imaging affects the surgical remission rates of Cushing’s disease. During the process of this review, we expect that we will be able to calculate an estimate of the remission rate using standard preoperative MRI only. In addition, by investigating sources of heterogeneity in outcomes (threshold for biochemical remission, for example) using statistical adjustments during meta-analysis, we may be able to discover predictors of surgical outcomes.

The results of this review will inform the directions and decisions regarding Cushing’s disease on three levels:

1. **Clinical** - inform best practices and guidelines regarding the surgical investigations and management of Cushing’s disease.
2. **Resources and management** - whether there is sufficient evidence for non-standard imaging modalities to merit resource reallocation, or at least a health economic analysis (the latter being outside the scope of this review)
3. **Research** - whether there are any promising avenues to be explored with regards to imaging for Cushing’s disease, or whether realignment of research priorities towards other areas might be more fruitful.

We plan to disseminate the results of this review through presentations at international conferences, and through peer-reviewed publications in established journals, with the strength of the evidence summarised according to the GRADE approach ^14^.

## Data Availability

Protocol - No data

